# Development and Validation of a Nomogram for Predicting Postoperative Lower Extremity Deep Vein Thrombosis in Patients with Traumatic Spinal Fractures

**DOI:** 10.1101/2025.10.05.25337375

**Authors:** Guangxu Fu, Yong Wang, Dengwu Tan, Zhen Zhang, Xiaoyan Yu

**Affiliations:** Department of Orthopedic Surgery, The People’s Hospital of Lichuan City, Enshi Tujia and miao Autonomous Prefecture, Hubei, China; Department of Anesthesiology, The People’s Hospital of Lichuan City, Enshi Tujia and miao Autonomous Prefecture, Hubei, China

**Keywords:** Spinal fractures, Deep vein thrombosis, Risk prediction model, Postoperative complications, Nomogram

## Abstract

**Background:** Patients undergoing surgery for traumatic spinal fractures face a substantially elevated risk of postoperative lower extremity deep vein thrombosis (DVT). While generic risk assessment tools exist, a purpose-built model integrating spine-specific and readily available preoperative predictors is lacking. This study aimed to develop and internally validate a novel predictive model for this specific complication.

**Methods:** This retrospective cohort study analyzed data from 1,676 patients who underwent surgery for traumatic spinal fractures at a single center. All patients received standardized DVT surveillance. The cohort was randomly split into training (70%) and testing (30%) sets. Univariate and multivariable logistic regression with stepwise selection were used to identify independent predictors from 29 candidate variables. Model performance was evaluated by its discriminative ability (area under the curve, AUC), calibration (calibration curves and Hosmer-Lemeshow test), and clinical utility (decision curve analysis, DCA). A nomogram was constructed for clinical use.

**Results:** The incidence of postoperative DVT was 14.26% (239/1,676). Six independent preoperative predictors were identified: prolonged bed rest > 72 hours (adjusted odds ratio [aOR] = 5.208), pre-existing lower extremity vascular disease (aOR = 2.938), elevated D-dimer (aOR = 1.582), elevated fibrinogen (aOR = 1.434), severe neurological impairment (ASIA grade A/B), and advanced age (aOR = 1.019). The model demonstrated robust discrimination (AUC: 0.891 training, 0.885 testing) and excellent calibration (Hosmer-Lemeshow p > 0.7), with high sensitivity (90.5– 91.2%) and moderate specificity (74.3–74.5%). Decision curve analysis confirmed its clinical utility across a wide range of threshold probabilities.

**Conclusion:** We developed and validated a parsimonious and clinically practical prediction model for postoperative DVT in traumatic spinal fracture patients. This tool, which leverages six preoperatively accessible variables, facilitates individualized risk stratification and could guide the implementation of targeted prophylactic strategies to improve patient outcomes.

## Introduction

Traumatic spinal fractures represent a major global public health challenge, with an estimated annual incidence of approximately 8.6 million cases, frequently resulting in long-term disability and substantial healthcare expenditures [1,2]. These injuries, typically caused by high-energy mechanisms such as motor vehicle collisions or falls, often require surgical intervention to restore spinal stability and create conditions conducive to neurological recovery [3,4]. The perioperative period, however, is marked by a significantly elevated risk of thrombotic complications [5]. Postoperative lower extremity deep vein thrombosis poses a significant clinical challenge in this patient population. It is critical to note that the threat of DVT is substantially present even prior to surgery, as evidenced by a reported preoperative DVT incidence of 14.45% in patients with high-energy thoracolumbar fractures [6]. Furthermore, during the postoperative phase, the incidence of DVT remains high at 18.91% despite the implementation of standardized low-molecular-weight heparin-based prophylaxis protocols [7]. These findings clearly indicate that while current conventional preventive strategies are necessary, they are remain inadequate, thereby emphasizing the pressing need for precise identification and stratification of high-risk individuals.

The clinical implications of DVT are profound and far-reaching, serving not only as the direct cause of fatal pulmonary embolism but also as the origin of post-thrombotic syndrome, which manifests as chronic limb swelling, pain, and ulceration, severely compromising patient rehabilitation and quality of life [8,9]. The basic pathophysiology underpinning this significant thrombotic tendency is the simultaneous activation of Virchow’s triad by both the original trauma and the following surgery. Venous stasis occurs as a result of limb paralysis caused by spinal cord damage or the forced immobilization required for spinal protection, which eliminates the calf muscle pump and substantially inhibits venous return in the lower extremities. Endothelial injury is caused not only directly by fracture fragments and surgical manipulation but also indirectly by a trauma-induced systemic inflammatory response, which promotes broad endothelial dysfunction [10,11]. Simultaneously, a systemic hypercoagulable state develops as tissue damage and surgery serve as significant stressors. This process triggers an acute-phase response that is characterized by the activation of coagulation factors, increased platelet reactivity, and suppression of the fibrinolytic system, all of which contribute to a prothrombotic environment [12].

In clinical practice, risk assessment often relies on generic models like the Caprini score. While these tools provide a valuable framework for Venous Thromboembolism (VTE) risk stratification in broad surgical populations, their performance may be suboptimal in specific, high-risk cohorts like traumatic spine fracture patients [13,14]. A key limitation is their failure to adequately incorporate disease-specific determinants of risk. The American Spinal Injury Association (ASIA) Impairment Scale, a gold-standard metric for neurological function that is a powerful surrogate for the degree of venous stasis and neurogenic hypercoagulability, is not systematically integrated [15]. Furthermore, while readily available preoperative biomarkers hold significant prognostic value, they are underutilized in existing prediction paradigms. Elevated levels of D-dimer, a fibrin degradation product indicating activated coagulation and fibrinolysis, and fibrinogen, a key acute-phase reactant and substrate for clot formation, have been independently associated with VTE risk in trauma and orthopedic populations [16,17]. Nevertheless, their combined predictive power, potentially refined as a ratio, has not been fully leveraged in a purpose-built model for spine surgery.

This critical gap underscores the pressing need for a specialized predictive tool that is both parsimonious and precisely calibrated to the unique pathophysiology of traumatic spinal fracture patients. Therefore, the primary objective of this study was to develop and internally validate a novel clinical prediction model that integrates key demographic, injury-specific, and biomarker data to accurately stratify the risk of postoperative DVT. We aimed to create an accessible nomogram to guide personalized prophylactic strategies, with the ultimate goal of improving outcomes in this vulnerable population.

## 2. Materials and methods

### 2.1 Study design and population

This retrospective cohort study was conducted using data from the electronic health records of the People’s Hospital of Lichuan City, covering patients admitted from January 2020 to March 2025. The authors did not have access to any personally identifiable information at any stage of the research. The study aimed to develop and internally validate a predictive model for postoperative lower extremity DVT in patients undergoing surgery for traumatic spinal fractures. The study protocol was approved by the Institutional Review Board of the hospital (Approval No.: 2025003). The requirement for informed consent was waived because the study followed the Declaration of Helsinki and used a retrospective design with anonymized data.

We included patients aged 18 and above who had a radiologically verified acute traumatic fracture of the cervical, thoracic, or lumbar spine and had open reduction and internal fixation. Furthermore, were required to have undergone comprehensive postoperative screening for lower extremity DVT using venous duplex ultrasonography within the first week of recovery and had medical records with > 80% completeness for all key characteristics. Exclusion criteria included: (1) a preoperative diagnosis of DVT or pulmonary embolism; (2) pre-existing severe coagulopathy or ongoing long-term therapeutic anticoagulation; (3) active malignancy or documented autoimmune systemic diseases; (4) concomitant pelvic or lower extremity fractures requiring surgical intervention; and (5) incomplete essential data or loss to follow-up prior to the primary outcome assessment.

A standardized protocol for DVT surveillance and prophylaxis was implemented throughout the study period. DVT diagnosis was objectively confirmed by board-certified radiologists using compression ultrasonography or, in equivocal cases, computed tomography venography. The surveillance protocol included preoperative D-dimer testing and baseline ultrasound. Postoperatively, all patients underwent daily clinical assessments for DVT signs, with scheduled follow-up ultrasounds on postoperative days 1, 3, and 7. Perioperative thromboprophylaxis followed an institutional protocol based on Caprini risk assessment. Low-risk patients (Caprini score ≤ 3) received mechanical prophylaxis with intermittent pneumatic compression devices, while moderate-to-high-risk patients (Caprini score ≥ 4) received combined mechanical prophylaxis and pharmacological anticoagulation, typically with low-molecular-weight heparin (e.g., enoxaparin 4000 IU subcutaneously once daily). Anticoagulation was initiated 6-12 hours postoperatively and continued for ≥ 14 days, or until 48 hours after significant independent ambulation.

The final cohort consisted of 1,676 patients, including 239 (14.2%) with postoperative DVT. With 29 candidate predictors considered, the events per variable (EPV) ratio was 8.24. While slightly below the heuristic of EPV ≥ 10, this ratio remains acceptable given the application of robust internal validation techniques to mitigate overfitting.

### 2.2 Collection of relevant variables

Data were systematically abstracted by trained research staff using a standardized electronic case report form. The collected variables, postulated to influence DVT risk based on existing literature, were organized into four domains: demographic and baseline characteristics, pre-existing comorbidities, perioperative and injury-related factors, and preoperative laboratory parameters. Demographic and baseline data included gender, age, body mass index (BMI), smoking status (categorized as current smoker or non-smoker), and alcohol consumption (categorized as regular drinker [> 210 g of pure alcohol per week] or non-drinker). This threshold is derived from the UK Chief Medical Officers’ guideline, where 210 g of alcohol approximates 14 standard UK units (1 unit = 8 g ethanol). Documented pre-existing comorbidities included coronary artery disease, hypertension, diabetes mellitus, cerebrovascular disease, chronic obstructive pulmonary disease (COPD) or pulmonary fibrosis, and lower extremity vascular disease. Perioperative and injury-related variables encompassed the location of fractures (cervical, thoracic, lumbar), the mechanism of injury (categorized as low-energy or high-energy), and neurologic status evaluated using the American Spinal Injury Association (ASIA) Impairment Scale. Additional factors included surgical methods (classified as internal fixation or decompression combined with internal fixation), total operative time (in minutes), estimated intraoperative blood loss (in milliliters), the receipt of any perioperative allogeneic blood transfusion, and preoperative bed rest time (measured in hours from hospital admission to the commencement of surgery). All preoperative laboratory parameters were measured from venous blood samples collected within 24 hours prior to surgery and included D-dimer, fibrinogen (FIB), prothrombin time (PT), activated partial thromboplastin time (APTT), platelet count (PLT), serum albumin (ALB), hemoglobin (Hb), C-reactive protein (CRP), and white blood cell count (WBC).

### 2.3 Statistical Analysis

Statistical analyses were conducted using SPSS (Version 26.0), R software (Version 4.3.1), and Zstats software (www.zstats.net), a dedicated platform for clinical statistical analysis. Continuous variables were assessed for normality with the Shapiro-Wilk test. Data presentation varied based on distribution: normally distributed variables are reported as mean ± standard deviation and compared using independent Student’s t-tests; non-normally distributed variables are reported as median with interquartile range (IQR) and compared using the Mann-Whitney U test. Categorical variables are summarized as frequencies and percentages, with group comparisons performed using either Pearson’s chi-square test or Fisher’s exact test, as appropriate.

The identification of independent risk factors and development of the predictive model followed a structured process. First, univariate logistic regression was performed, and variables with a significance level of p < 0.1 were retained as candidates for multivariable analysis. To ensure the stability of the subsequent multivariable model, multicollinearity among these candidate variables was assessed using the generalized variance inflation factor (GVIF), which is appropriate for models containing categorical predictors. A GVIF value below 2.5 for all variables indicated the absence of substantial multicollinearity, and thus no variables required elimination.A multivariable logistic regression model was then fitted using backward stepwise selection based on the likelihood ratio test, with the goal of deriving a parsimonious final model. Variables with a p-value < 0.05 in this final model were considered significant independent predictors.

The final model was graphically depicted as a nomogram created with R’s rms package. Model validation followed a two-pronged internal validation strategy. Initially, the dataset was randomly partitioned into a training cohort (70%) for model development and a testing cohort (30%) for validation. Subsequently, bootstrap resampling with 1,000 repetitions was performed on the complete dataset to generate bias-corrected performance estimates. Model discrimination was quantified by the area under the receiver operating characteristic curve (AUC). Calibration performance, reflecting the agreement between predicted probabilities and observed outcomes, was evaluated using calibration curves and the Hosmer-Lemeshow goodness-of-fit test (where p > 0.05 indicates adequate calibration). Additional performance metrics, including sensitivity, specificity, accuracy, positive predictive value, and negative predictive value, were calculated from the classification tables. The clinical net benefit of implementing the model across various probability thresholds was assessed using decision curve analysis (DCA).

## 3. Result

### 3.1 Baseline demographics and characteristics

Among the 1,676 patients in this study, 239 (14.26%) developed postoperative lower extremity DVT, while 1,437 (85.74%) did not. Patients with DVT were significantly older than those in the Non-DVT group (median age 60.0 [IQR 49.0-67.5] vs 51.0 [41.0-62.0]; P < 0.001). The DVT group had a higher percentage of injuries with ASIA grade A (28.45% vs 4.80%; P < 0.001) and grade B (18.41% vs 6.26%; P < 0.001), indicating significant neurological impairment. Lower extremity vascular disease was substantially more prevalent in the DVT group (25.94% vs 8.91%; P < 0.001). Preoperative bed rest time exceeding 72 hours was significantly higher in the DVT group, with rates of 64.44% compared to 26.93% (P < 0.001). Laboratory parameters differed significantly, with the DVT group exhibiting elevated D-dimer levels, with a median of 5.30 mg/L (IQR 2.95-7.65) versus 1.60 mg/L (1.00-2.80) in the control group (P < 0.001). Additionally, FIB levels were also higher in the DVT group, with a median of 4.70 g/L (IQR 4.00-5.30) compared to 4.20 g/L (3.50-4.80) in the control group (P < 0.001). SSignificant differences were also observed in surgical factors. The total operative time was significantly longer in the DVT group, with a median of 144.0 minutes (IQR 116.5-178.0) compared to 133.0 minutes (109.0-164.0) in the control group (P = 0.002). Furthermore, the DVT group experienced greater intraoperative blood loss, with a median of 250.0 mL (IQR 160.0-320.0) versus 230.0 mL (140.0-290.0) in the control group (P = 0.048).

No statistically significant differences were identified between the two groups concerning gender, BMI, smoking or alcohol consumption status, comorbidities (including coronary artery disease, hypertension, diabetes, cerebrovascular disease, and COPD/pulmonary fibrosis), fracture location, injury mechanism, surgical technique, blood transfusion requirements, or other laboratory parameters such as PT, APTT, PLT, ALB, Hb, CRP, and WBC (all P > 0.05). The baseline characteristics of the entire cohort are comprehensively presented in Table 1. Furthermore, the dataset was randomly divided into training (n = 1,175) and testing (n = 501) sets. As shown in Table S1, all baseline variables were well-balanced between the two sets, with no significant differences (all P > 0.05), confirming the appropriateness of the split for model development and validation. The baseline characteristics of the training set, stratified by DVT status, are provided in Table S2, which similarly reflected the significant predictors identified in the overall cohort.

**Table 1.** Baseline Characteristics.

| Variables | Total (n = 1676) | Non-DVT (n = 1437) | DVT (n = 239) | Statistic | P |
| --- | --- | --- | --- | --- | --- |
| Gender, n(%) | | | | $\chi^2=0.283$ | 0.595 |
| Femal | 696 (41.53) | 593 (41.27) | 103 (43.10) |  |  |
| Male | 980 (58.47) | 844 (58.73) | 136 (56.90) |  |  |
| Age, M (Q<sub>1</sub>, Q<sub>3</sub>) | 52.00 (42.00, 63.00) | 51.00 (41.00, 62.00) | 60.00 (49.00, 67.50) | $Z=-6.532$ | <.001 |
| BMI (Kg/m<sup>2</sup>), n(%) | | | | $\chi^2=4.426$ | 0.219 |
| ≤18.4 | 113 (6.74) | 91 (6.33) | 22 (9.21) |  |  |
| 18.5–23.9 | 843 (50.30) | 721 (50.17) | 122 (51.05) |  |  |
| 24.0–27.9 | 518 (30.91) | 445 (30.97) | 73 (30.54) |  |  |
| ≥28.0 | 202 (12.05) | 180 (12.53) | 22 (9.21) |  |  |
| Smoking, n(%) | | | | $\chi^2=0.606$ | 0.436 |
| No | 1295 (77.27) | 1115 (77.59) | 180 (75.31) |  |  |
| Yes | 381 (22.73) | 322 (22.41) | 59 (24.69) |  |  |
| Drinking, n(%) | | | | $\chi^2=0.080$ | 0.777 |
| No | 1316 (78.52) | 1130 (78.64) | 186 (77.82) |  |  |
| Yes | 360 (21.48) | 307 (21.36) | 53 (22.18) |  |  |
| Coronary artery disease, n(%) | | | | $\chi^2=1.191$ | 0.275 |
| No | 1541 (91.95) | 1317 (91.65) | 224 (93.72) |  |  |
| Yes | 135 (8.05) | 120 (8.35) | 15 (6.28) |  |  |
| Hypertension, n(%) | | | | $\chi^2=0.197$ | 0.657 |
| No | 1193 (71.18) | 1020 (70.98) | 173 (72.38) |  |  |
| Yes | 483 (28.82) | 417 (29.02) | 66 (27.62) |  |  |
| Diabetes, n(%) | | | | $\chi^2=0.002$ | 0.967 |
| No | 1446 (86.28) | 1240 (86.29) | 206 (86.19) |  |  |
| Yes | 230 (13.72) | 197 (13.71) | 33 (13.81) |  |  |
| Cerebrovascular disease, n(%) | | | | $\chi^2=0.281$ | 0.596 |
| No | 1583 (94.45) | 1359 (94.57) | 224 (93.72) |  |  |
| Yes | 93 (5.55) | 78 (5.43) | 15 (6.28) |  |  |
| COPD or pulmonary fibrosis, n(%) | | | | $\chi^2=1.368$ | 0.242 |
| No | 1608 (95.94) | 1382 (96.17) | 226 (94.56) |  |  |
| Yes | 68 (4.06) | 55 (3.83) | 13 (5.44) |  |  |
| Lower extremity vascular disease, n(%) | | | | $\chi^2=59.155$ | <.001 |
| No | 1486 (88.66) | 1309 (91.09) | 177 (74.06) |  |  |
| Yes | 190 (11.34) | 128 (8.91) | 62 (25.94) |  |  |
| Fracture location, n(%) | | | | $\chi^2=1.924$ | 0.382 |
| Cervical | 336 (20.05) | 284 (19.76) | 52 (21.76) |  |  |
| Thoracic | 499 (29.77) | 422 (29.37) | 77 (32.22) |  |  |
| Lumbar | 841 (50.18) | 731 (50.87) | 110 (46.03) |  |  |
| Injury mechanism, n(%) | | | | $\chi^2=3.417$ | 0.065 |
| Low-energy injury | 646 (38.54) | 541 (37.65) | 105 (43.93) |  |  |
| High-energy injury | 1030 (61.46) | 896 (62.35) | 134 (56.07) |  |  |
| ASIA grade, n(%) | | | | $\chi^2=228.419$ | <.001 |
| A | 137 (8.17) | 69 (4.80) | 68 (28.45) |  |  |
| B | 134 (8.00) | 90 (6.26) | 44 (18.41) |  |  |
| C | 178 (10.62) | 143 (9.95) | 35 (14.64) |  |  |
| D | 466 (27.80) | 434 (30.20) | 32 (13.39) |  |  |
| E | 761 (45.41) | 701 (48.78) | 60 (25.10) |  |  |
| Surgical Methods, n(%) | | | | $\chi^2=0.205$ | 0.650 |
| Internal Fixation | 1164 (69.45) | 1001 (69.66) | 163 (68.20) |  |  |
| Decompression + Internal Fixation | 512 (30.55) | 436 (30.34) | 76 (31.80) |  |  |
| Blood transfusion, n(%) | | | | $\chi^2=0.124$ | 0.724 |
| No | 1305 (77.86) | 1121 (78.01) | 184 (76.99) |  |  |
| Yes | 371 (22.14) | 316 (21.99) | 55 (23.01) |  |  |
| Total Operative Time (min) , M (Q<sub>1</sub>, Q<sub>3</sub>) | 134.00 (110.00, 166.00) | 133.00 (109.00, 164.00) | 144.00 (116.50, 178.00) | Z=-3.066 | **0.002** |
| Intraoperative blood loss (ml), M (Q<sub>1</sub>, Q<sub>3</sub>) | 230.00 (147.50, 300.00) | 230.00 (140.00, 290.00) | 250.00 (160.00, 320.00) | Z=-1.979 | **0.048** |
| Preoperative bed rest time, n(%) | | | | $\chi^2=131.854$ | <.001 |
| ≤ 72 h | 1135 (67.72) | 1050 (73.07) | 85 (35.56) |  |  |
| > 72 h | 541 (32.28) | 387 (26.93) | 154 (64.44) |  |  |
| D-Dimer(mg/L), M (Q<sub>1</sub>, Q<sub>3</sub>) | 1.80 (1.00, 3.40) | 1.60 (1.00, 2.80) | 5.30 (2.95, 7.65) | Z=-16.366 | <.001 |
| FIB (g/L), M (Q<sub>1</sub>, Q<sub>3</sub>) | 4.20 (3.60, 4.90) | 4.20 (3.50, 4.80) | 4.70 (4.00, 5.30) | Z=-6.374 | <.001 |
| PT (s), M (Q<sub>1</sub>, Q<sub>3</sub>) | 12.30 (11.10, 13.40) | 12.30 (11.10, 13.40) | 12.30 (11.20, 13.35) | Z=-0.471 | 0.638 |
| APTT (s), M (Q<sub>1</sub>, Q<sub>3</sub>) | 29.65 (23.80, 35.90) | 29.80 (24.00, 35.80) | 29.00 (22.90, 36.75) | Z=-0.515 | 0.606 |
| PLT (10<sup>9</sup>/L), M (Q<sub>1</sub>, Q<sub>3</sub>) | 303.93 (226.72, 378.36) | 303.89 (227.05, 378.27) | 306.93 (225.69, 377.13) | Z=-0.196 | 0.844 |
| ALB (g/L), M (Q<sub>1</sub>, Q<sub>3</sub>) | 34.87 (31.43, 38.53) | 34.87 (31.40, 38.69) | 34.80 (31.51, 37.67) | Z=-1.202 | 0.229 |
| Hb (g/L), M (Q<sub>1</sub>, Q<sub>3</sub>) | 108.30 (97.10, 120.53) | 108.80 (97.40, 120.70) | 105.70 (95.15, 119.65) | Z=-1.473 | 0.141 |
| CRP (mg/L), M (Q<sub>1</sub>, Q<sub>3</sub>) | 26.35 (15.41, 37.31) | 26.19 (15.50, 37.26) | 27.39 (13.50, 37.50) | Z=-0.238 | 0.812 |
| WBC (10<sup>9</sup>/L), M (Q<sub>1</sub>, Q<sub>3</sub>) | 11.00 (9.10, 13.00) | 11.00 (9.10, 12.90) | 11.20 (9.20, 13.10) | Z=-0.833 | 0.405 |

Z: Mann-Whitney U test, $\chi^2$ : Chi-square test, M: Median, Q<sub>1</sub>: 1st Quartile, Q<sub>3</sub>: 3rd Quartile, BMI: body mass index, COPD: chronic obstructive pulmonary disease, ASIA grade: American Spinal Injury Association grade, FIB: fibrinogen, PT: prothrombin time, APTT: activated partial thromboplastin time, PLT: platelet count, ALB: serum albumin, Hb: hemoglobin, CRP: C-reactive protein, WBC: white blood cell count.

### 3.2 Independent risk factors for Lower Extremity DVT Following Traumatic Spinal Fracture Surgery

Univariate logistic regression was performed to identify potential risk factors associated with postoperative lower extremity DVT (Table 2). Variables yielding a P < 0.1 in this initial screening were selected as candidates for the multivariable analysis. These included age, lower extremity vascular disease, ASIA grade, preoperative bed rest time, D-dimer, FIB, and BMI categories.

**Table 2.**
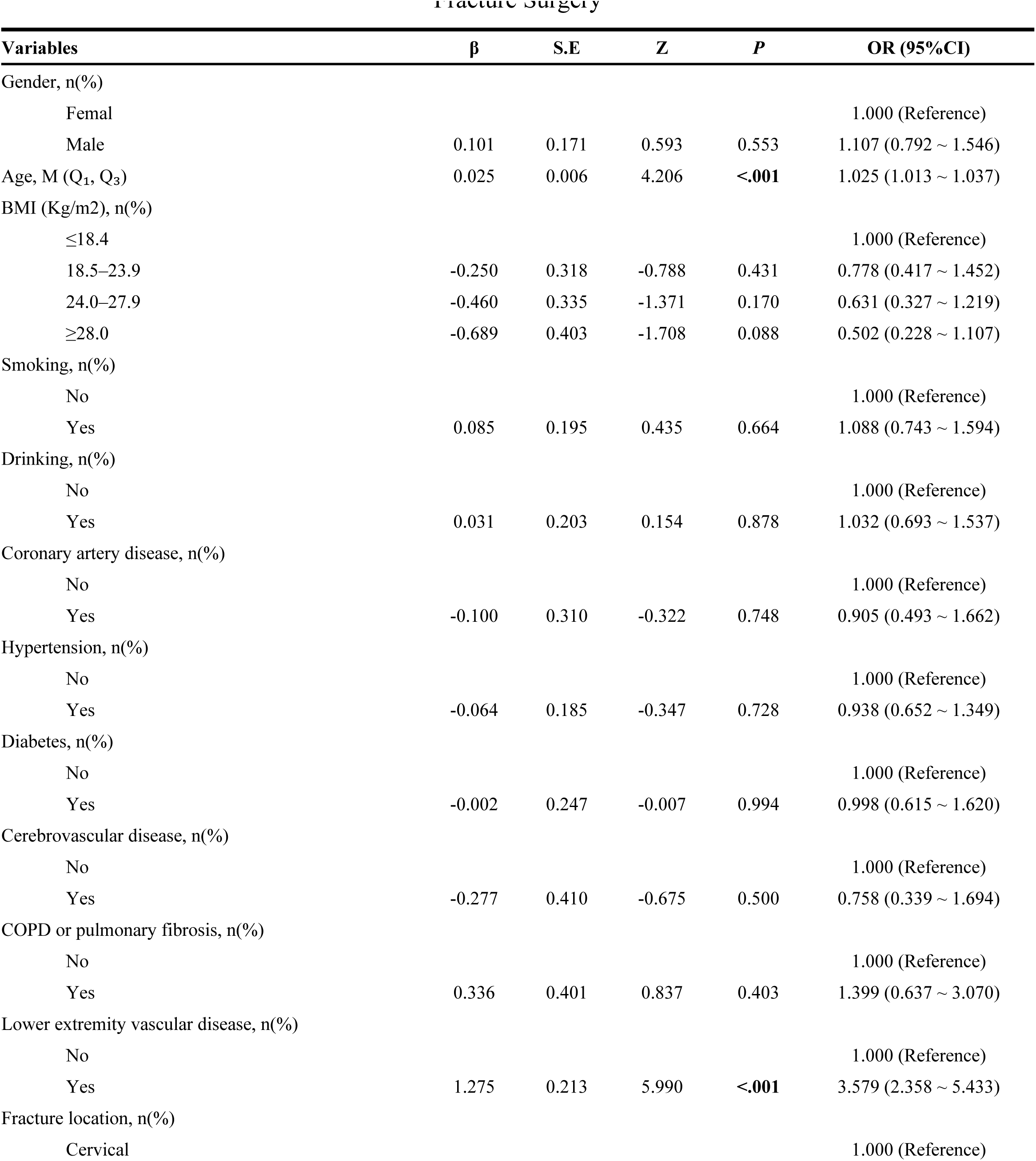
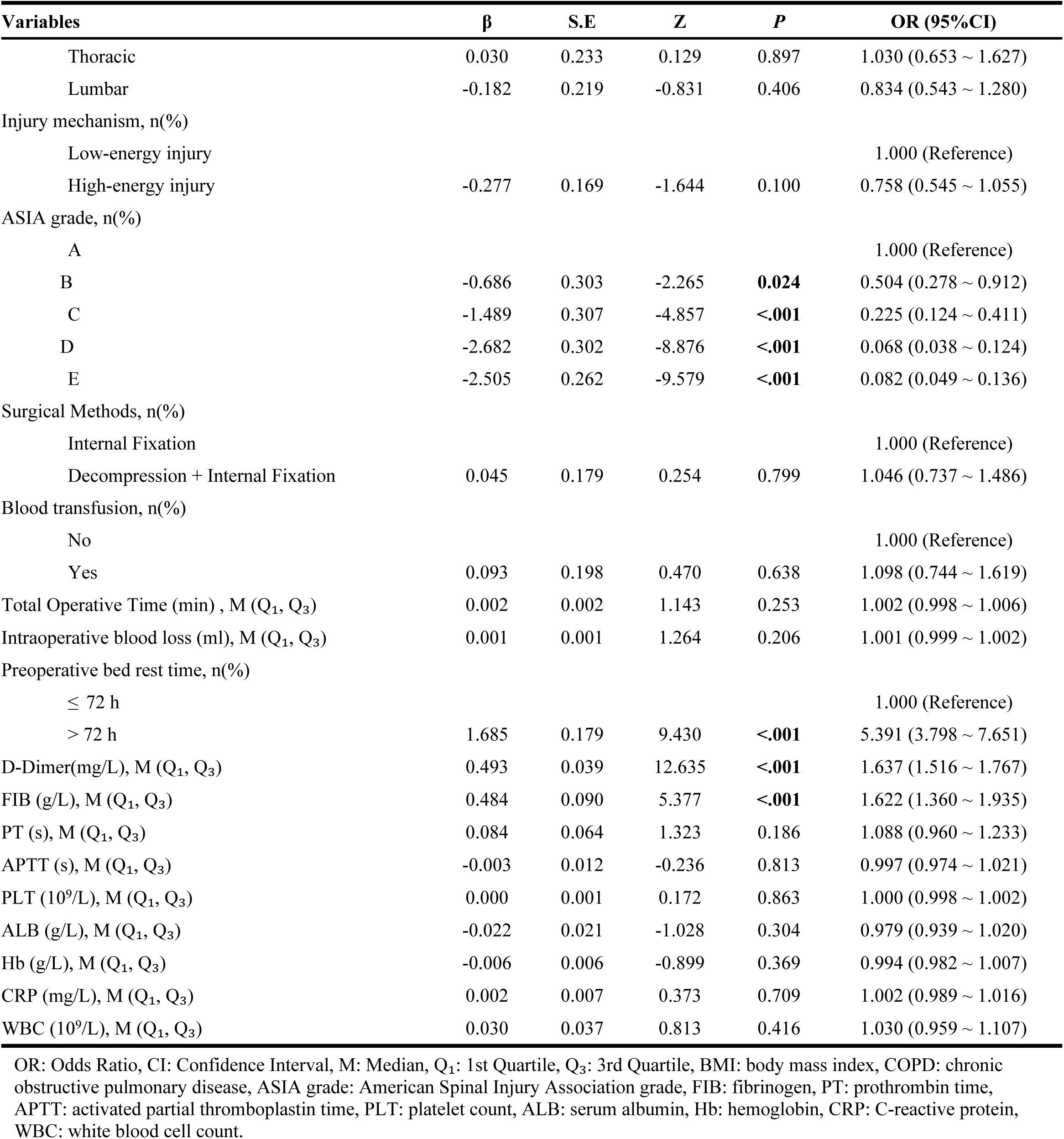
Univariate analysis of risk factors for Lower Extremity DVT Following Traumatic Spinal Fracture Surgery.

To construct a robust and parsimonious predictive model, the candidate variables were first entered into a full multivariable logistic regression model. Prior to model interpretation, we assessed multicollinearity among these predictors using the GVIF. All GVIF values were below 1.1, indicating an absence of significant multicollinearity and confirming that all candidate variables could be retained for further model refinement. Subsequently, we employed a stepwise variable selection procedure based on the Akaike Information Criterion (AIC) to optimize model fit while avoiding overfitting. This process excluded the BMI categories due to their non-significant contribution to the model, resulting in a final model with six independent predictors.

The final multivariable logistic regression model, which was optimized using a stepwise selection method, demonstrated six independent factors that were significantly correlated with the onset of postoperative lower extremities DVT (Table 3). The primary risk factors identified included a preoperative bed rest duration beyond 72 hours (adjusted odds ratio [aOR] = 5.208, 95% confidence interval [CI]: 3.319-8.171; P < 0.001) and the existence of lower extremity vascular disease (aOR = 2.938, 95% CI: 1.641-5.258; P < 0.001). Furthermore, laboratory biomarkers indicative of hypercoagulability were found to be significantly associated, with each unit rise in D-dimer level (aOR = 1.582, 95% CI: 1.448-1.729; P < 0.001) and FIB (aOR = 1.434, 95% CI: 1.138-1.807; P = 0.002) associated with an elevated risk. The model demonstrated a robust, graded inverse correlation between neurological function and the probability of DVT. Utilizing ASIA grade A (the most severe injury) as the benchmark, the likelihood of DVT diminished steadily with decreasing severity: grade B (aOR = 0.332, P = 0.007), grade C (aOR = 0.234, P < 0.001), grade D (aOR = 0.070, P < 0.001), and grade E (aOR = 0.097, P < 0.001). Advanced age was also independently associated with an increased risk (aOR = 1.019 per year, 95% CI: 1.003-1.035; P = 0.018). The model demonstrated a favorable fit, as indicated by a significant reduction in deviance from the null model (residual deviation: 561.52 with 1165 degrees of freedom). The final AIC value of 581.52 for the stepwise model was lower than that of the full model (AIC: 586), which confirms the enhanced parsimony of the final model while maintaining its explanatory power.

**Table 3.**
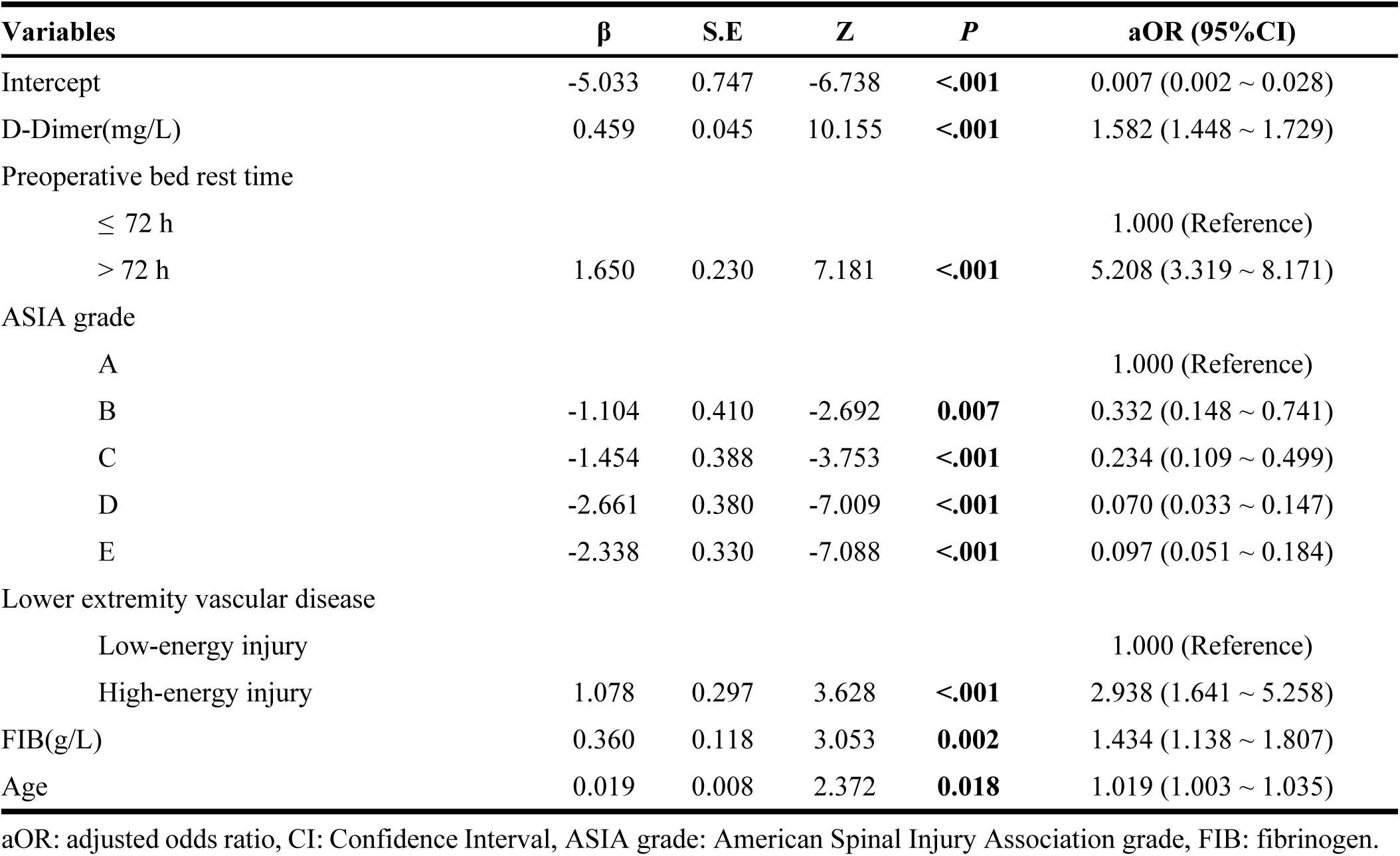
Multivariate Logistic Regression Analysis of Risk Factors for Lower Extremity DVT.

| Variables | $\beta$ | S.E | Z | P | aOR (95%CI) |
| --- | --- | --- | --- | --- | --- |
| Intercept | -5.033 | 0.747 | -6.738 | <.001 | 0.007 (0.002 ~ 0.028) |
| D-Dimer(mg/L) | 0.459 | 0.045 | 10.155 | <.001 | 1.582 (1.448 ~ 1.729) |
| Preoperative bed rest time |  |  |  |  |  |
| $\leq 72$ h | | | | | 1.000 (Reference) |
| $> 72$ h | 1.650 | 0.230 | 7.181 | <.001 | 5.208 (3.319 ~ 8.171) |
| ASIA grade |  |  |  |  |  |
| A |  |  |  |  | 1.000 (Reference) |
| B | -1.104 | 0.410 | -2.692 | 0.007 | 0.332 (0.148 ~ 0.741) |
| C | -1.454 | 0.388 | -3.753 | <.001 | 0.234 (0.109 ~ 0.499) |
| D | -2.661 | 0.380 | -7.009 | <.001 | 0.070 (0.033 ~ 0.147) |
| E | -2.338 | 0.330 | -7.088 | <.001 | 0.097 (0.051 ~ 0.184) |
| Lower extremity vascular disease |  |  |  |  |  |
| Low-energy injury |  |  |  |  | 1.000 (Reference) |
| High-energy injury | 1.078 | 0.297 | 3.628 | <.001 | 2.938 (1.641 ~ 5.258) |
| FIB(g/L) | 0.360 | 0.118 | 3.053 | 0.002 | 1.434 (1.138 ~ 1.807) |
| Age | 0.019 | 0.008 | 2.372 | 0.018 | 1.019 (1.003 ~ 1.035) |

aOR: adjusted odds ratio, CI: Confidence Interval, ASIA grade: American Spinal Injury Association grade, FIB: fibrinogen.

### 3.3 Nomogram construction and validation

Based on the multivariable logistic regression analysis, a nomogram was developed to predict the risk of postoperative lower extremity DVT in patients with traumatic spinal fractures, incorporating significant independent predictors (Fig 1A). The model was constructed using the training cohort (n = 1,175) and validated both internally via bootstrapping with 1,000 repetitions and externally in an independent testing cohort (n = 501). The nomogram demonstrated strong discriminative performance, with area under the curve (AUC) values of 0.891 (95% CI: 0.862-0.919) in the training set and 0.885 (95% CI: 0.849-0.921) in the testing set (Fig 1B-C). Sensitivity was high in both cohorts: 91.2% (95% CI: 89.4-92.9%) in the training set and 90.5% (95% CI: 88.4-92.6%) in the testing set. Specificity was 74.3% (95% CI: 67.6-80.9%) and 74.5% (95% CI: 66.5-82.5%) in the training and testing sets, respectively (Table S3). The model achieved an overall accuracy of 88.8% (95% CI: 86.8-90.5%) in the training set and 87.0% (95% CI: 84.5-89.2%) in the testing set. Positive predictive values were 95.5% and 95.2%, while negative predictive values were 58.2% and 62.5% in the training and testing sets, respectively.

**Fig 1.**
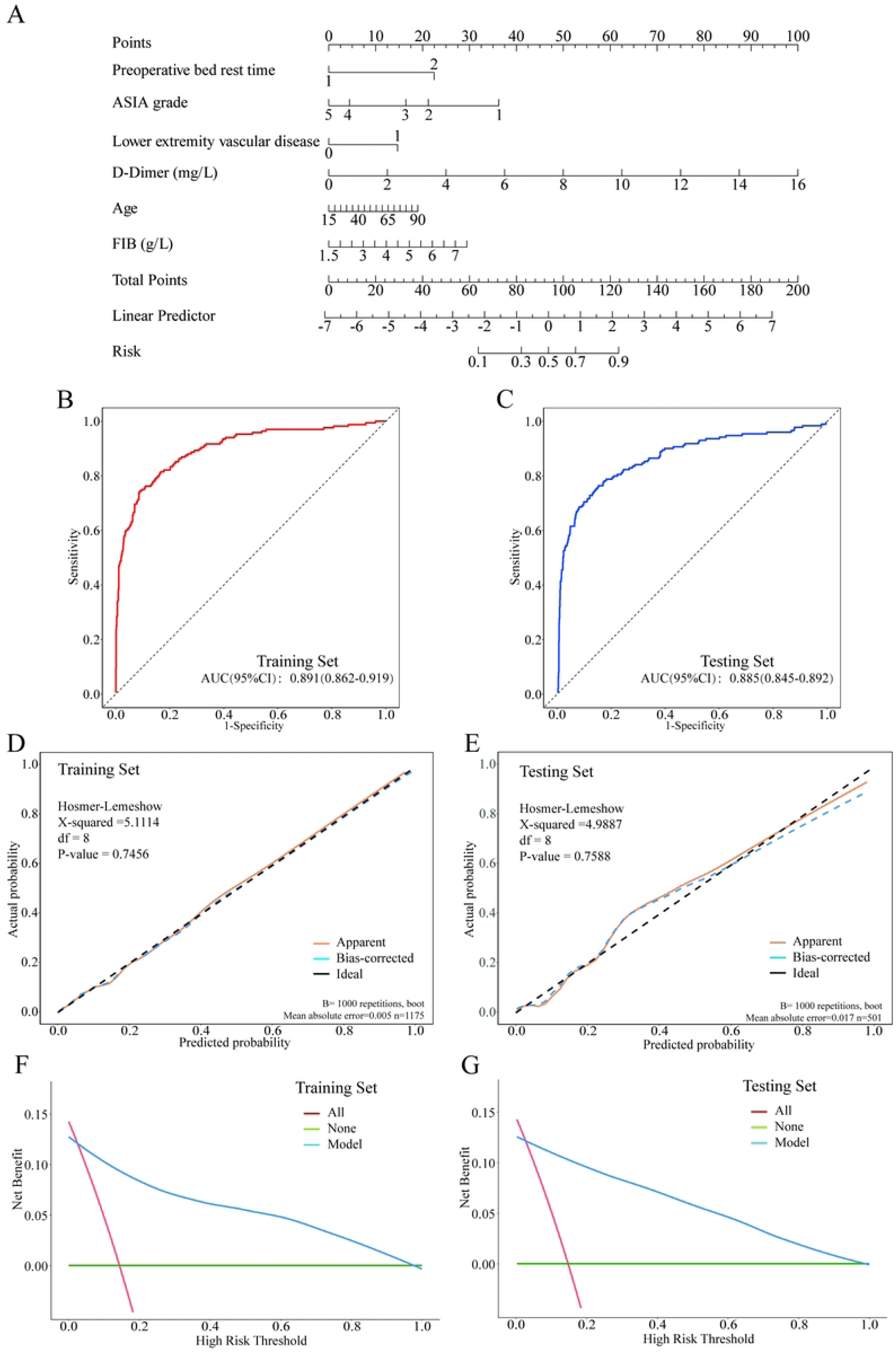
Illustrates the development and validation of the DVT risk prediction nomogram. Depicted are: (A) the nomogram; ROC curves for (B) training and (C) testing sets; calibration curves for (D) training and (E) testing sets; and decision curve analyses for (F) training and (G) testing sets.

The model’s calibration was evaluated using the Hosmer-Lemeshow test, indicating good agreement between predicted and observed probabilities in both the training (χ² = 5.1114, df = 8, p = 0.7456) and testing (χ² = 4.9887, df = 8, p = 0.7588) datasets (Fig 1D-E). Bootstrap validation improved model robustness, with mean absolute errors of 0.005 and 0.017 for the training and testing datasets, respectively. DCA revealed that the nomogram provided superior net benefit across a wide range of clinically relevant threshold probabilities compared to the “treat all” or “treat none” strategies (Fig 1F-G), underscoring its potential utility for individualized risk assessment and clinical decision-making. The resulting nomogram incorporates key factors and demonstrates excellent discrimination, calibration, and net benefit in both the training and validation cohorts, supporting its potential clinical utility.

## 4. Discussion

This study developed and internally validated a predictive model for postoperative lower extremity DVT in patients with traumatic spinal fractures, based on a large retrospective cohort. The final model incorporated six preoperatively available predictors, forming a concise risk stratification tool. It demonstrated excellent discriminatory performance in the validation set, with an area under the curve of 0.885, alongside satisfactory calibration. Decision curve analysis further indicated that the model provides significant clinical net benefit across a wide range of threshold probabilities, underscoring its potential for translation into clinical practice.

Mandatory bed rest following spinal trauma often induces a severe hypodynamic state in the lower extremities. The weakening or loss of the calf muscle pump function significantly reduces venous blood flow velocity, leading to marked venous stasis, which represents the primary pathophysiological step in DVT development [18,19]. Studies have consistently shown a strong correlation between the duration of immobilization and DVT risk, with a significant increase observed particularly after exceeding the critical threshold of 3 days [20]. Furthermore, the risk escalates with prolonged bed rest. Early mobilization, when feasible and safe, has been demonstrated to positively impact the prevention and prognosis of DVT [21,22]. Our findings quantitatively confirm preoperative bed rest time as the strongest predictor of DVT, aligning with observations by Kerwin et al., who reported a daily increase in venous thromboembolism risk with prolonged immobilization following major trauma [23].

More importantly, the underlying pathology extends beyond mere physical reduction in blood flow. Research indicates that venous stasis creates a localized hypoxic microenvironment. Recent investigations reveal that low shear stress alone can mechanically activate the HIF-1α pathway even under normoxic conditions [24]. A 2025 in vitro model of trauma-induced endotheliopathy provides clear evidence that endothelial cells activate the HIF-1α pathway in a complicated traumatic milieu. Concurrently, the study found that thrombomodulin was downregulated and shed, while tissue factor expression was upregulated in these cells. This “molecular switch” directly causes the endothelium to change from an anticoagulant to a procoagulant phenotype, compromising its intrinsic anticoagulant function [25]. Thus, immobilization synergistically enhances thrombogenesis via two pathways: “venous stasis” and “endothelial injury.”

In this study, D-dimer and fibrinogen were established as two key and independent biological predictors for postoperative DVT following traumatic spinal fracture surgery. Their synergistic action accurately reflects the core coexisting pathophysiological processes in postoperative patients: secondary hyperfibrinolysis and a systemic hypercoagulable state. Specifically, elevated D-dimer signifies the generation and degradation of cross-linked fibrin, serving as direct evidence of active thrombosis and dissolution in vivo [26,27]. Surgical trauma is a recognized potent activator of the coagulation-fibrinolysis system. Notably, recent research has shifted focus towards the dynamic trajectory of D-dimer levels rather than relying solely on single measurements. A study involving gastrointestinal surgery patients introduced the concept of “7-day cumulative D-dimer exposure,” finding that patients who developed DVT maintained persistently higher postoperative D-dimer levels, exhibiting a “high-baseline, slow-decline” pattern compared to the “rapid decline” seen in non-DVT patients. This cumulative exposure was an independent predictor of DVT [28]. This suggests that dynamic monitoring of D-dimer may hold superior predictive value compared to a single measurement in spinal trauma patients. Meanwhile, fibrinogen, as a liver-synthesized acute-phase protein, contributes to thrombosis not only by increasing the substrate concentration for clot formation and blood viscosity but also by potentially forming denser, more rigid fibrin clots that are more resistant to fibrinolysis [29]. Omics research has revealed that fibrinogen, along with platelet parameters, forms a significantly elevated “PLT cluster” in DVT patients. This cluster shows positive correlation with specific metabolites, offering a novel metabolic perspective on the complex role of fibrinogen in DVT pathogenesis [30]. The combined use of these two biomarkers provides greater value than either one used alone. Research indicates that integrating D-dimer and fibrinogen with additional laboratory data could significantly enhance the predictive capabilities of standard risk assessment models, such as the Caprini score [31]. A risk prediction model tailored for fracture patients that included both D-dimer and fibrinogen demonstrated exceptional predictive efficacy, reinforcing the notion that their combination enhances the algorithm’s ability to identify high-risk individuals [32].

A distinctive strength of our model lies in its formal integration of neurological status as measured by the ASIA Impairment Scale, which demonstrating a robust inverse correlation between neurological deficiency and DVT risk. This finding is consistent with a previous large-scale clinical investigation, which found that patients with thoracic spinal cord injuries and ASIA grade A/B deficits had a roughly fourfold higher incidence of VTE than others [33]. The underlying mechanisms extend far beyond simple flaccid paralysis and stasis, potentially involving dysautonomia affecting venous tone. Severe spinal cord injury triggers a systemic inflammatory response, with experimental studies confirming significant upregulation of inflammatory cytokines like IL-6 at the injury site [34]. Concurrently, trauma itself, as a potent stressor, can induce an acquired hypercoagulable state, wherein functional inhibition of anticoagulant pathways, including the protein C system, is a key component [35]. Furthermore, thoracic injuries can lead to autonomic dysregulation, potentially exacerbating venous stasis by impairing venous tone [36]. These multifactorial mechanisms act in concert to drive a systemic hypercoagulable state, establishing neurological deficit as a core driver of thrombogenesis.

Furthermore, the model encompasses foundational patient susceptibility factors. Pre-existing lower extremity vascular disease implies baseline endothelial dysfunction or venous insufficiency, significantly lowering the threshold for de novo thrombosis under the combined stress of trauma and surgery. Reviews on superficial venous thrombosis emphasize that lower extremity venous disease and DVT share common risk factors, explaining why a substantial proportion of patients with superficial vein thrombosis also present with concomitant DVT [37]. Advanced age, a consistent risk factor across thrombotic diseases, reflects the confluence of age-related physiological declines: reduced venous compliance, diminished physiological reserve, and a well-documented propensity towards a prothrombotic haemostatic balance. Large-scale epidemiological studies have confirmed the association between age and VTE risk at the population level, while recent mechanistic research suggests that age-related coagulation changes might exacerbate the accumulation risk of certain anticoagulants [38], providing additional pathophysiological rationale for age as an independent risk factor.

The compelling performance metrics of our nomogram support its potential for integration into routine clinical practice. First, the development and validation of the prediction model were based on a substantial, well-characterized cohort of patients undergoing surgery for traumatic spinal fractures, enhancing the statistical reliability and potential generalizability of our findings. Second, the model is inherently clinically practical, as it relies exclusively on preoperatively accessible variables, facilitating its potential integration into routine clinical workflows for early risk stratification. Third, we employed rigorous internal validation techniques, including data splitting and bootstrap resampling, which provide robust evidence for the model’s discriminative ability, calibration, and clinical utility, thereby mitigating concerns of overfitting. Finally, the identified predictors are not merely statistical associations but are firmly grounded in the established pathophysiology of Virchow’s triad, lending biological plausibility and coherence to our model.

Notwithstanding these strengths, several limitations warrant careful consideration. he retrospective, single-center design poses risks of unmeasured confounding and may limit the generalizability of our findings. While our DVT surveillance protocol was standardized, the reliance on duplex ultrasonography—especially for identifying asymptomatic distal thrombi—may have resulted in an underestimation of the actual DVT incidence. Additionally, we were unable to account for specific variations in pharmacological thromboprophylaxis regimens, which could indicate residual confounding. Ultimately, confirming the clinical impact of this model requires external validation in multi-center prospective cohorts and, ideally, an interventional trial to assess its implementation on patient outcomes.

In conclusion, we have developed and rigorously validated a clinically applicable prediction model for postoperative lower extremity DVT following traumatic spinal fracture surgery. This parsimonious tool, which integrates six readily available preoperative predictors, demonstrates strong predictive performance and the potential to facilitate individualized, risk-adapted thromboprophylaxis. Future research should focus on the external validation and clinical implementation of this model to ultimately improve outcomes in this vulnerable patient population.

## Abbreviations

AIC: Akaike Information Criterion
ALB: Albumin
APTT: Activated Partial Thromboplastin Time
ASIA: American Spinal Injury Association
AUC: Area Under the Curve
BMI: Body Mass Index
COPD: Chronic Obstructive Pulmonary Disease
CRP: C-Reactive Protein
DCA: Decision Curve Analysis
DVT: Deep Vein Thrombosis
FIB: Fibrinogen
GVIF: Generalized Variance Inflation Factor
Hb: Hemoglobin
IQR: Interquartile Range
aOR: Adjusted Odds Ratio
PLT: Platelet Count
PT: Prothrombin Time
WBC: White Blood Cell Count

## Data Availability

The datasets generated and analyzed during this study are not publicly available due to institutional data governance policies. However, de-identified data may be made available by the corresponding author upon reasonable request and with approval from the relevant ethics committee.

## Acknowledgements

The authors would like to thank the patients in our study.

## Author contributions

Conceptualization: Guangxu Fu, Xiaoyan Yu

Data Curation: Guangxu Fu, Yong Wang, Dengwu Tan Formal Analysis: Guangxu Fu

Funding Acquisition: Xiaoyan Yu

Investigation: Guangxu Fu, Yong Wang, Dengwu Tan

Methodology: Guangxu Fu, Xiaoyan Yu Project Administration: Xiaoyan Yu Resources: Guangxu Fu

Software: Guangxu Fu, Zhen Zhang Supervision: Xiaoyan Yu

Validation: Guangxu Fu, Xiaoyan Yu Visualization: Guangxu Fu

Writing-Original Draft Preparation: Guangxu Fu

Writing-Review & Editing: Xiaoyan Yu, Yong Wang, Dengwu Tan

## Declarations

### Ethics approval and consent to participate

The study was conducted in accordance with the Declaration of Helsinki and received approval from the Medical Ethics Committee of the People’s Hospital of Lichuan City (approval number: 2025003). Prior to analysis, all data were anonymised, and patient consent was deemed unnecessary due to the retrospective nature of the study.

### Consent for publication

Not applicable.

### Clinical trial number

Not applicable.

